# Demographic and Outcome Characteristics of Children Hospitalized with Acute COVID-19 versus Multisystem Inflammatory Syndrome in Children in Canada

**DOI:** 10.1101/2022.08.18.22278939

**Authors:** Daniel S. Farrar, Charlotte Moore Hepburn, Olivier Drouin, Tala El Tal, Marie-Paule Morin, Roberta A. Berard, Melanie King, Melanie Laffin Thibodeau, Elie Haddad, Rosie Scuccimarri, Rae S. M. Yeung, Fatima Kakkar, Shaun K. Morris, the Canadian Paediatric Surveillance Program COVID-19 Study Team

**Affiliations:** Centre for Global Child Health, The Hospital for Sick Children, Toronto, Canada; Division of Paediatric Medicine, The Hospital for Sick Children, Toronto, Canada; Institute of Health Policy, Management and Evaluation, University of Toronto, Toronto, Canada; Division of Rheumatology, Department of Pediatrics, Children’s Hospital at London Health Sciences Centre, London, Canada; Division of General Pediatrics, Department of Pediatrics, CHU Sainte-Justine, Montreal, Canada; Department of Social and Preventive Medicine, School of Public Health, Université de Montréal, Montréal, Canada; Division of Rheumatology, Department of Pediatrics, The Hospital for Sick Children, University of Toronto, Canada; Division of Paediatric Rheumatology-Immunology, CHU Sainte-Justine, Department of Pediatrics, University of Montreal, Montreal, Canada; Canadian Paediatric Surveillance Program, Canadian Paediatric Society, Ottawa, Canada; Division of Pediatric Rheumatology, Montreal Children’s Hospital and McGill University Health Centre, Montreal, Canada; Cell Biology Program, The Hospital for Sick Children, Toronto, Canada; Department of Immunology and Institute of Medical Science, University of Toronto, Toronto, Canada; Division of Infectious Diseases, CHU Sainte-Justine, Montreal, Canada; Division of Infectious Diseases, The Hospital for Sick Children, Toronto, Canada; Department of Pediatrics, Temerty Faculty of Medicine, University of Toronto, Toronto, Canada; Clinical Public Health, Dalla Lana School of Public Health, University of Toronto, Toronto, Canada

**Keywords:** COVID-19, multisystem inflammatory syndrome in children, hospitalizations, resource use, intensive care

## Abstract

Direct comparisons of pediatric hospitalizations for acute COVID-19 and multisystem inflammatory syndrome in children (MIS-C) can inform health system planning. While there were more hospitalizations and deaths from acute COVID-19 amongst Canadian children between March 2020–May 2021, MIS-C cases were more severe, requiring more intensive care and vasopressor support.

## INTRODUCTION

Along with hospitalization for acute COVID-19, multisystem inflammatory syndrome in children (MIS-C) has emerged as a serious yet infrequent complication of pediatric SARS-CoV-2 infection. To date, few studies have directly compared characteristics and outcomes associated with these two diagnoses.(1,2) Case series describing MIS-C indicate higher proportions of severe disease relative to acute COVID-19, despite much lower incidence of MIS-C in the community compared to SARS-CoV-2 infection.(3,4) Differences in the associated use of hospital resources (e.g. ventilation or hemodynamic support requiring intensive care) between these two disease entities are not well known, and may have implications for future pediatric pandemic planning. We aimed to describe the absolute and relative hospital burden of acute pediatric COVID-19 infection and MIS-C during the first fifteen months of the pandemic in Canada, prior to the emergence of the Omicron variant and approval of SARS-CoV-2 vaccines for use in children.

## METHODS

We conducted a national prospective study via the Canadian Paediatric Surveillance Program (CPSP) from March 2020–May 2021. The CPSP is a public health surveillance network which includes >2800 paediatricians across Canada, who voluntarily reported incident cases to this study on a weekly basis. Cases of children <18 years old and hospitalized with acute SARS-CoV-2 infection or pediatric inflammatory multisystem syndrome (PIMS) were eligible to be reported. While cases were reported based on a surveillance definition of PIMS, we applied a post-hoc case definition of MIS-C according to the World Health Organization.(5) By definition, all patients with MIS-C had a documented linkage to SARS-CoV-2 (i.e. positive polymerase chain reaction, rapid antigen, or serology test, or a close contact with microbiologically confirmed SARS-CoV-2). For all SARS-CoV-2 hospitalizations, the reporting physician indicated whether the hospitalization was due to acute COVID-19 or if incidental infection was identified upon routine screening; this was confirmed by dual adjudication by the study team to ensure consistency. We therefore compared two mutually exclusive groups for this analysis: children hospitalized with acute COVID-19 versus children hospitalized with MIS-C. Further details regarding the study design are described elsewhere, and surveillance definitions are available at https://cpsp.cps.ca/surveillance/study-etude/covid-19.(6,7) Ethics approval was obtained at Health Canada-PHAC (REB #2020-002P), the Hospital for Sick Children (REB #1000070001), and the Centre Hospitalier Universitaire Sainte-Justine (conducted as a multicenter study in the province of Quebec; IRB #MP-21-2021-2901).

Baseline characteristics and severity outcomes were ascertained for both case definitions using the same case report form. Severity outcomes included requirements for supplemental oxygen (either low-flow oxygen or high-flow nasal cannula), ventilation (either non-invasive, conventional mechanical, or high-frequency oscillatory ventilation), vasopressors, pediatric intensive care unit (PICU) admissions, or death. Characteristics were summarized using medians, interquartile ranges (IQR), frequencies, and percentages. Frequencies between one and four were reported as ‘<5’ while some larger frequencies were presented as ranges to prevent back-calculation, in accordance with CPSP privacy policy. Adjusted risk differences (aRD) were calculated to identify factors associated with each diagnosis, adjusting for age, sex, presence of ≥1 comorbid conditions, and the timing of hospitalization (defined as five three-month periods from March 2020–May 2021). Differences in continuous variables (i.e. age and PICU length of stay) were assessed using Wilcoxon rank-sum tests. The temporal lag (in weeks) between all Canadian SARS-CoV-2 case counts (ascertained from the Public Health Agency of Canada(8)) and hospitalizations reported to CPSP were assessed using Spearman’s rank correlation coefficient. P-values <0.05 were considered statistically significant. Analyses were conducted in Stata v17.0.

## RESULTS

Overall, 330 children hospitalized with acute COVID-19 and 208 hospitalized with MIS-C were reported during the surveillance period (Table 1). The median age among acute COVID-19 patients (1.9 years; IQR 0.1–13.3) was significantly younger than those with MIS-C (8.1 years; IQR 4.2–11.6; p<0.001). More children aged <1 year were hospitalized with acute COVID-19 than with MIS-C (aRD 43.4%; 95% CI 37.7–49.1), while more children aged 5–11 years were hospitalized with MIS-C than acute COVID-19 (aRD 38.9%; 95% CI 31.0–46.9). Chronic comorbid conditions were more common amongst acute COVID-19 patients (43.0% vs. 15.9% with MIS-C; aRD 38.0%; 95% CI 31.0–45.1).

**Table 1.** Characteristics of children hospitalized with acute COVID-19 and MIS-C.

| Characteristic | Diagnosis |  | aRD<sup>1</sup>, % | 95% CI | P value |
| --- | --- | --- | --- | --- | --- |
|  | COVID-19 | MIS-C |  |  |  |
| **Total hospitalizations, N<sup>2</sup>** | 330 | 208 | --- | --- | --- |
| **Age (years), median (IQR)<sup>3</sup>** | 1.9 (0.1–13.3) | 8.1 (4.2–11.6) | --- | --- | <0.001 |
| **Age (years), n (%)** |  |  |  |  |  |
| <1 | 140 (42.4) | 9 (4.3) | 43.4 | 37.7 to 49.1 | <0.001 |
| 1–4 | 68 (20.6) | 52 (25.0) | -2.9 | -10.4 to 4.6 | 0.45 |
| 5–11 | 30 (9.1) | 98 (47.1) | -38.9 | -46.9 to -31.0 | <0.001 |
| 12–17 | 92 (27.9) | 49 (23.6) | -1.6 | -9.6 to 6.4 | 0.69 |
| **Sex, n (%)** |  |  |  |  |  |
| Female | 145 (43.9) | 76 (36.5) | 6.2 | -4.9 to 17.3 | 0.27 |
| Male | 185 (56.1) | 132 (63.5) | -6.2 | -17.3 to 4.9 | 0.27 |
| **Comorbid conditions<sup>4</sup>, n (%)** |  |  |  |  |  |
| None/Unknown | 188 (57.0) | 175 (84.1) | -38.0 | -45.1 to -31.0 | <0.001 |
| ≥1 | 142 (43.0) | 33 (15.9) | 38.0 | 31.0 to 45.1 | <0.001 |
| **Population group, n (%)** |  |  |  |  |  |
| White | 75 (22.7) | 63 (30.3) | -12.4 | -22.8 to -2.0 | 0.02 |
| South Asian | 40 (12.1) | 25 (12.0) | 1.4 | -6.0 to 8.7 | 0.71 |
| Arab/West Asian | 39 (11.8) | 16 (7.7) | 4.4 | -2.3 to 11.1 | 0.20 |
| Black | 39 (11.8) | 29 (13.9) | -2.2 | -9.5 to 5.1 | 0.56 |
| Indigenous | 28 (8.5) | 9 (4.3) | 4.8 | -0.6 to 10.3 | 0.08 |
| East/Southeast Asian | 18 (5.5) | 12 (5.8) | 3.0 | -2.2 to 8.1 | 0.26 |
| Latin American | 5 (1.5) | 18 (8.7) | -7.5 | -13.3 to -1.7 | 0.01 |
| Unknown | 92 (27.9) | 49 (23.6) | --- | --- | --- |

aRD = Adjusted risk difference; CI = Confidence interval; IQR = Interquartile range; MIS-C = Multisystem inflammatory syndrome in children
<sup>1</sup>All comparisons adjusted for age category, sex, comorbid conditions, and timing of hospitalization. Positive risk differences indicate a characteristic was more common for acute COVID-19 while negative risk differences indicate a characteristic was more common for MIS-C.
<sup>2</sup>Overall, 544 hospitalized children with SARS-CoV-2 infection were reported to the CPSP, among whom 330 children were admitted due to acute COVID-19; 493 hospitalized children were reported with suspected PIMS, among whom 208 children met the WHO definition of MIS-C.
<sup>3</sup>Continuous age was missing for four patients with acute COVID-19, though categorical age could still be determined.
<sup>4</sup>Comorbidities were reported as any of the following conditions: congenital heart disease, diabetes, gastrointestinal/liver disease, genetic/metabolic conditions, hematologic disorders, immunodeficiencies, malignancies, neurologic or neurodevelopmental conditions, obesity, pulmonary disease (i.e. asthma or other chronic lung disease), renal disease, rheumatologic/autoimmune disorders, tracheostomy, transplants, or other conditions.

PICU admission was required for 49.5% of MIS-C hospitalizations versus 18.2% of acute COVID-19 hospitalizations (aRD 20.3; 95% CI 9.9–30.8), though the proportion of children <5 years admitted to PICU was similar (19.7% vs. 15.4%; Table 2). However, the median length of PICU stay was one day greater for acute COVID-19 (4 days; IQR 2–7) than MIS-C (3 days; IQR 2–4; p=0.04). Vasopressor use was more common for MIS-C than acute COVID-19 at all ages (35.6% vs. 2.4%; aRD 23.1%; 95% CI 15.8–30.4). The proportion of all patients requiring supplemental oxygen and mechanical ventilation were similar (24.6% and 10.9% for acute COVID-19 vs. 30.3% and 9.6% for MIS-C, respectively). Five deaths due to acute COVID-19 were reported versus zero due to MIS-C.

**Table 2.** Outcomes of acute COVID-19 and MIS-C hospitalizations, overall and by age category.

| Outcome<sup>1</sup> | Diagnosis |  | aRD<sup>2</sup>, % | 95% CI | P value |
| --- | --- | --- | --- | --- | --- |
|  | COVID-19 | MIS-C |  |  |  |
| All hospitalizations |  |  |  |  |  |
| Supplemental oxygen | 81/330 (24.6) | 63/208 (30.3) | -5.6 | -15.2 to 3.9 | 0.25 |
| Ventilation | 36/330 (10.9) | 20/208 (9.6) | 1.7 | -4.9 to 8.2 | 0.62 |
| Vasopressors | 8/330 (2.4) | 74/208 (35.6) | -23.1 | -30.4 to -15.8 | <0.001 |
| PICU admission | 60/330 (18.2) | 103/208 (49.5) | -20.3 | -30.8 to -9.9 | <0.001 |
| PICU length of stay (days) | 4 (2–7) | 3 (2–4) | --- | --- | 0.04 |
| ECMO | 0/330 (0.0) | 0/208 (0.0) | --- | --- | --- |
| Death | 5/330 (1.5) | 0/208 (0.0) | --- | --- | --- |
| Age <5 years |  |  |  |  |  |
| Supplemental oxygen | 36/208 (17.3) | 10/61 (16.4) | -0.1 | -13.0 to 12.6 | 0.98 |
| Ventilation | 18–21/208 (8.7–10.1)<sup>3</sup> | <5/61 (<8.2) | 12.8 | 7.0 to 18.5 | <0.001 |
| Vasopressors | 5–7/208 (2.4–3.4)<sup>3</sup> | 8/61 (13.1) | -4.7 | -12.1 to 2.6 | 0.21 |
| PICU admission | 32/208 (15.4) | 12/61 (19.7) | 2.5 | -8.8 to 13.8 | 0.66 |
| PICU length of stay (days) | 4 (2–6) | 3 (1–3) | --- | --- | 0.02 |
| Age 5–11 years |  |  |  |  |  |
| Supplemental oxygen | 8/30 (26.7) | 37/98 (37.8) | -12.9 | -35.0 to 9.3 | 0.25 |
| Ventilation | <5/30 (<16.7) | 7–10/98 (7.1–10.2)<sup>3</sup> | -9.8 | -18.5 to -1.1 | 0.03 |
| Vasopressors | 0/30 (0.0) | 42/98 (42.9) | --- | --- | --- |
| PICU admission | 7/30 (23.3) | 61/98 (62.2) | -34.5 | -57.7 to -11.3 | 0.004 |
| PICU length of stay (days) | 2 (1–9) | 3 (2–4) | --- | --- | 0.68 |
| Age 12–17 years |  |  |  |  |  |
| Supplemental oxygen | 37/92 (40.2) | 16/49 (32.7) | -9.2 | -27.2 to 8.9 | 0.32 |
| Ventilation | 11–14/92 (12.0–15.2)<sup>3</sup> | 6–9/49 (12.2–18.4)<sup>3</sup> | -6.9 | -25.1 to 11.3 | 0.46 |
| Vasopressors | <5/92 (<5.4) | 24/49 (49.0) | -43.5 | -63.3 to -23.8 | <0.001 |
| PICU admission | 21/92 (22.8) | 30/49 (61.2) | -38.1 | -57.2 to -19.0 | <0.001 |
| PICU length of stay (days) | 6 (3–8) | 4 (2–5) | --- | --- | 0.11 |

aRD = Adjusted risk difference; CI = Confidence interval; ECMO = Extracorporeal membrane oxygenation; PICU = Pediatric intensive care unit; MIS-C = Multisystem inflammatory syndrome in children
<sup>1</sup>Descriptive statistics are presented as frequency and proportions, or medians and interquartile ranges.
<sup>2</sup>All comparisons adjusted for sex, comorbid conditions, and timing of hospitalization. Comparisons for all hospitalizations also adjust for age category. Positive risk differences indicate an outcome was more common for acute COVID-19 while negative risk differences indicate an outcome was more common for MIS-C.
<sup>3</sup>Frequencies and percentages are reported using ranges to prevent back-calculation of frequencies <5.

Acute pediatric COVID-19 hospitalization trends lagged behind all Canadian SARS-CoV-2 infection waves by one week (Spearman’s ρ 0.89), versus a lag of six weeks for MIS-C hospitalizations (Spearman’s ρ 0.82; Appendix Figure 1).

## DISCUSSION

Understanding the severity and associated in-hospital resource use required to manage acute pediatric COVID-19 and MIS-C is necessary to anticipate acute health system needs, and to make informed decisions regarding preventative measures including SARS-CoV-2 vaccination. Using national surveillance data from March 2020–May 2021, acute COVID-19 resulted in more pediatric hospitalizations and deaths and longer PICU stays, while MIS-C resulted in more PICU admissions and more frequent need for hemodynamic support. In this study, half of hospitalized MIS-C patients required intensive care, consistent with prior studies from the United Kingdom (44%) and United States (64%) during the first year of the pandemic.(9,10) While PICU admissions among MIS-C patients were often initiated due to shock, these patients were likely stabilized rapidly with immune modulation and vasopressor support. This may explain the shorter PICU stays relative to patients with acute COVID-19, who typically require PICU admission due to respiratory distress and/or exacerbation of chronic comorbid conditions (e.g. neurologic or respiratory disease). The higher vasopressor and intensive care requirements for MIS-C, and similar rates of respiratory support requirements, are consistent with U.S. data from Feldstein et al (2021).(1) A small number of acute COVID-19-related deaths were reported versus none due to MIS-C, in part due to complications from chronic comorbid conditions among children with severe COVID-19.(11)

Despite comparable absolute rates of overall in-hospital resource use, there were important age differences in disease severity and the ensuing strategies used to support these patients. Infants (i.e. <1 year old) rarely required hospitalization for MIS-C, presenting with lower rates of shock, coagulopathy, and myocarditis relative to older children.(7,12) Conversely, infants were the most commonly hospitalized age group for acute COVID-19, in part due to the routine practice of admitting febrile infants for investigations and empiric treatment. Meanwhile, the requirement for hemodynamic support among children ≥5 years with MIS-C (43–49%) likely led to the high proportion of PICU admission among this age group relative to acute COVID-19, in keeping with other published literature.(2)

While our study period ended prior to dominance of the Delta and Omicron lineages, these and future SARS-CoV-2 lineages may affect the relative in-hospital burden of acute COVID-19 infection and MIS-C. Studies from Denmark and Israel have found the incidence of MIS-C during Omicron waves fell to one-tenth that of prior waves, after accounting for vaccination status.(13,14) Declines in the proportion of MIS-C patients admitted to PICU have also been observed (e.g. 49% during Delta waves to 21% during Omicron waves in Israel(14)), though this may also reflect increased physician knowledge of MIS-C and refinement of supportive and treatment strategies. Uptake of pediatric and adolescent SARS-CoV-2 vaccines may also alter the relative in-hospital burden of disease, having shown effectiveness against both severe COVID-19 and MIS-C.(15)

There are several limitations to this study. First, the voluntary nature of CPSP reporting means that not all pediatric hospitalizations in Canada were identified. Moreover, the limited availability of molecular and serologic testing during the early pandemic likely resulted in some cases failing to meet the case definition of MIS-C. Data were also collected prior to emergence of the Delta and Omicron variants, and before implementation of pediatric and adolescent SARS-CoV-2 vaccine programs. PICU admission criteria may have differed by age, centre, and diagnosis. Nevertheless, this study provided a unique opportunity to compare children hospitalized for acute COVID-19 infection and MIS-C using data ascertained with the same surveillance methods, timeframe, and patient population.

Our findings suggest that both acute COVID-19 and MIS-C need to be considered when assessing the overall burden of SARS-CoV-2 in hospitalized children, and have implications for future pandemic planning with respect to hospital resource use. Given the high proportion of children requiring PICU support for MIS-C, in tandem with the limited number of specialized hospital beds, it is clear these resources need to be anticipated for future pandemic waves. Moreover, given the low overall rates of vaccination among children aged <12 years, awareness of disease severity from both acute COVID-19 and MIS-C may inform parents and policymakers in their decision-making regarding pediatric vaccines.

## Data Availability

De-identified data that underlie the results reported in this article (text, tables, and figure) and that abide by the privacy rules of the Canadian Paediatric Surveillance Program and the Public Health Agency of Canada can be made available to investigators whose secondary data analysis study protocol has been approved by an independent research ethics board.

## Abbreviations

(aRD): Adjusted risk difference
(CPSP): Canadian Paediatric Surveillance Program
(CI): confidence interval
(COVID-19): coronavirus disease 2019
(IQR): interquartile range
(MIS-C): multisystem inflammatory syndrome in children
(PHAC): Public Health Agency of Canada
(PICU): pediatric intensive care unit
(SARS-CoV-2): Severe acute respiratory disease coronavirus 2

## Supplemental Materials

**Appendix Figure 1.**
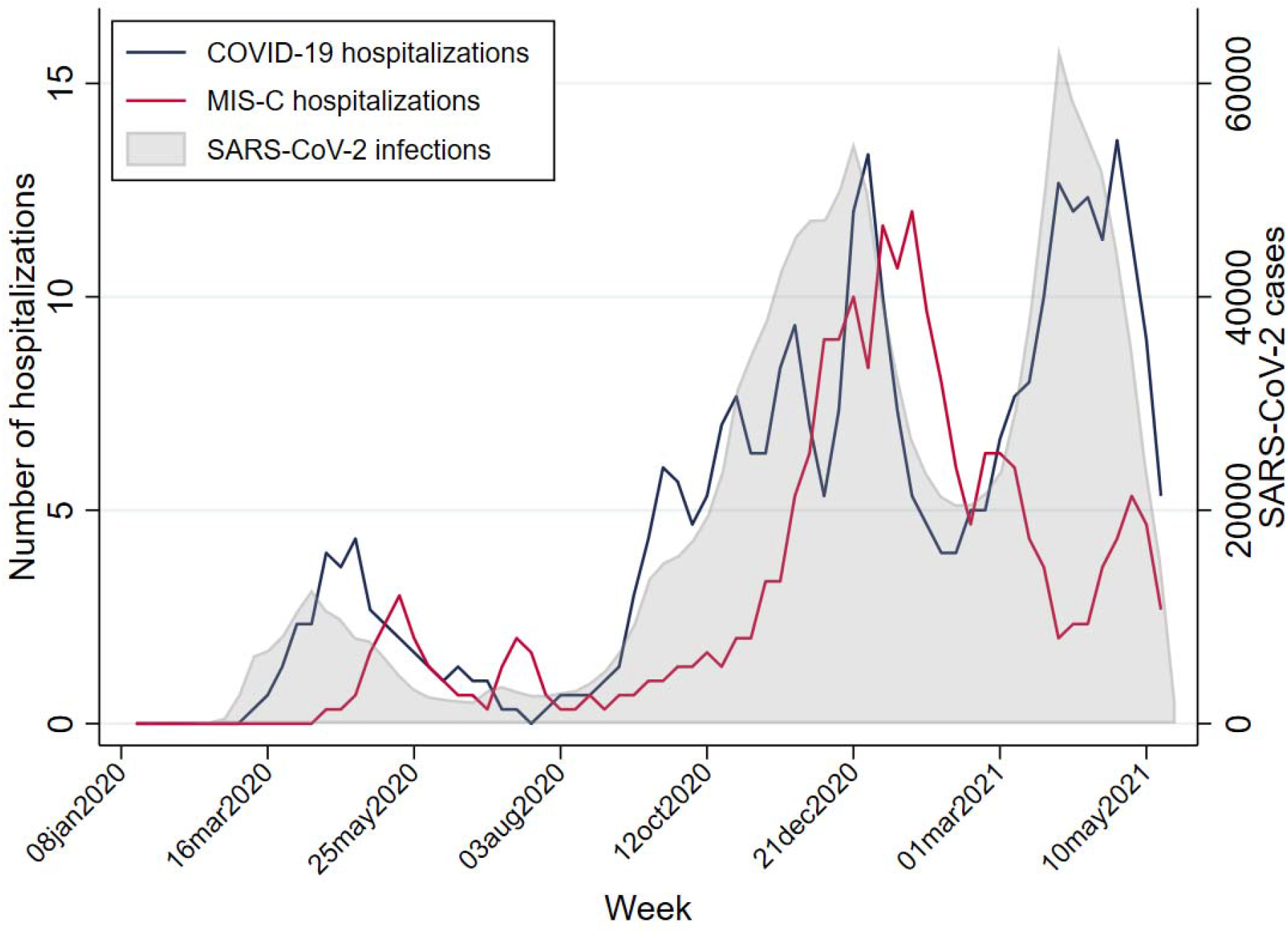
Time series of acute COVID-19 hospitalizations (<18 years), MIS-C hospitalizations (<18 years), and SARS-CoV-2 infections (all ages) across Canada from January 2020–May 2021. Data for COVID-19 and MIS-C hospitalizations represent the three-week moving average of cases included in this study. SARS-CoV-2 infections were ascertained from the Public Health Agency of Canada, available at https://health-infobase.canada.ca/covid-19/epidemiological-summary-covid-19-cases.html, and reflect the date of illness onset.

## CPSP COVID-19 Study Team (listed alphabetically)

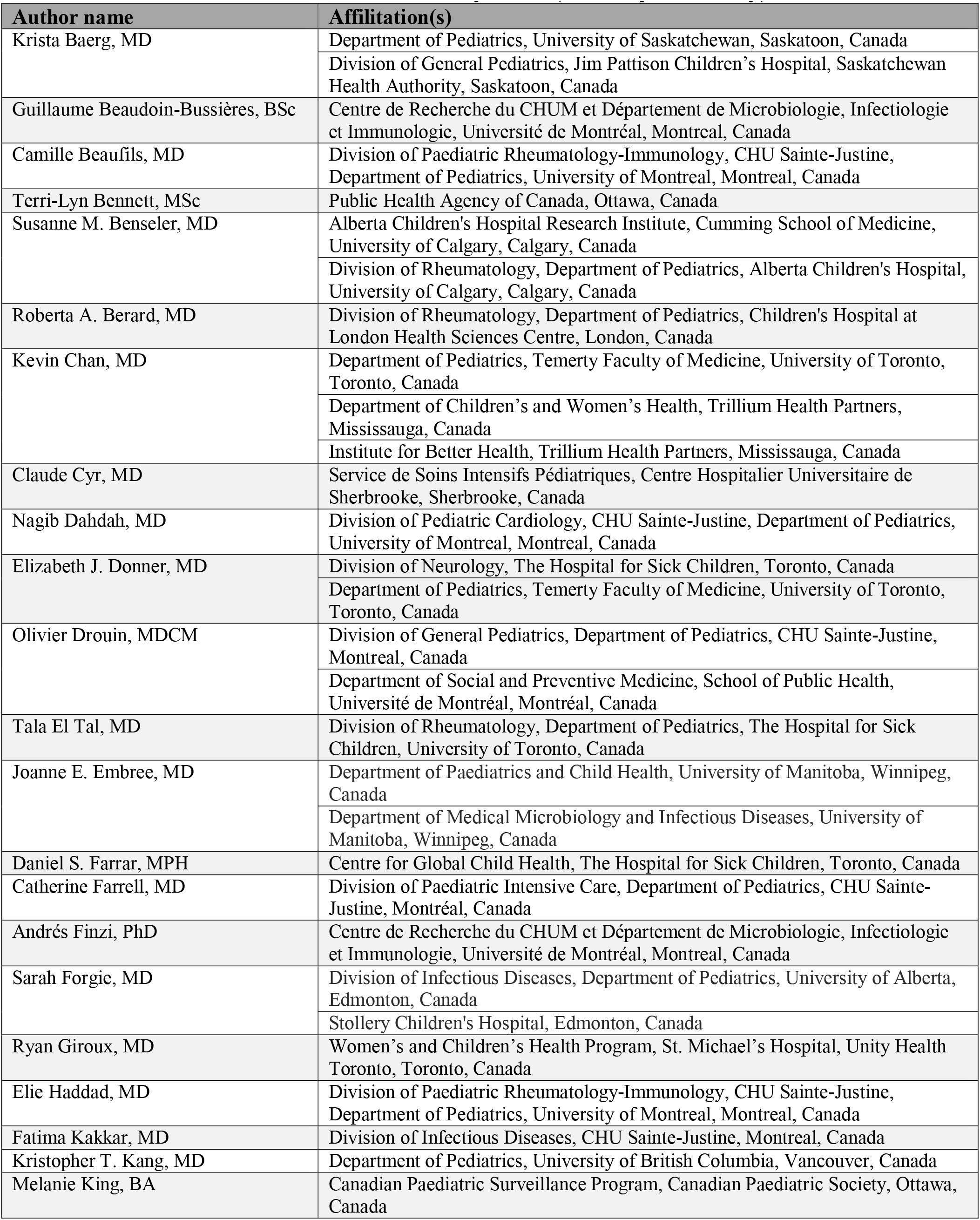
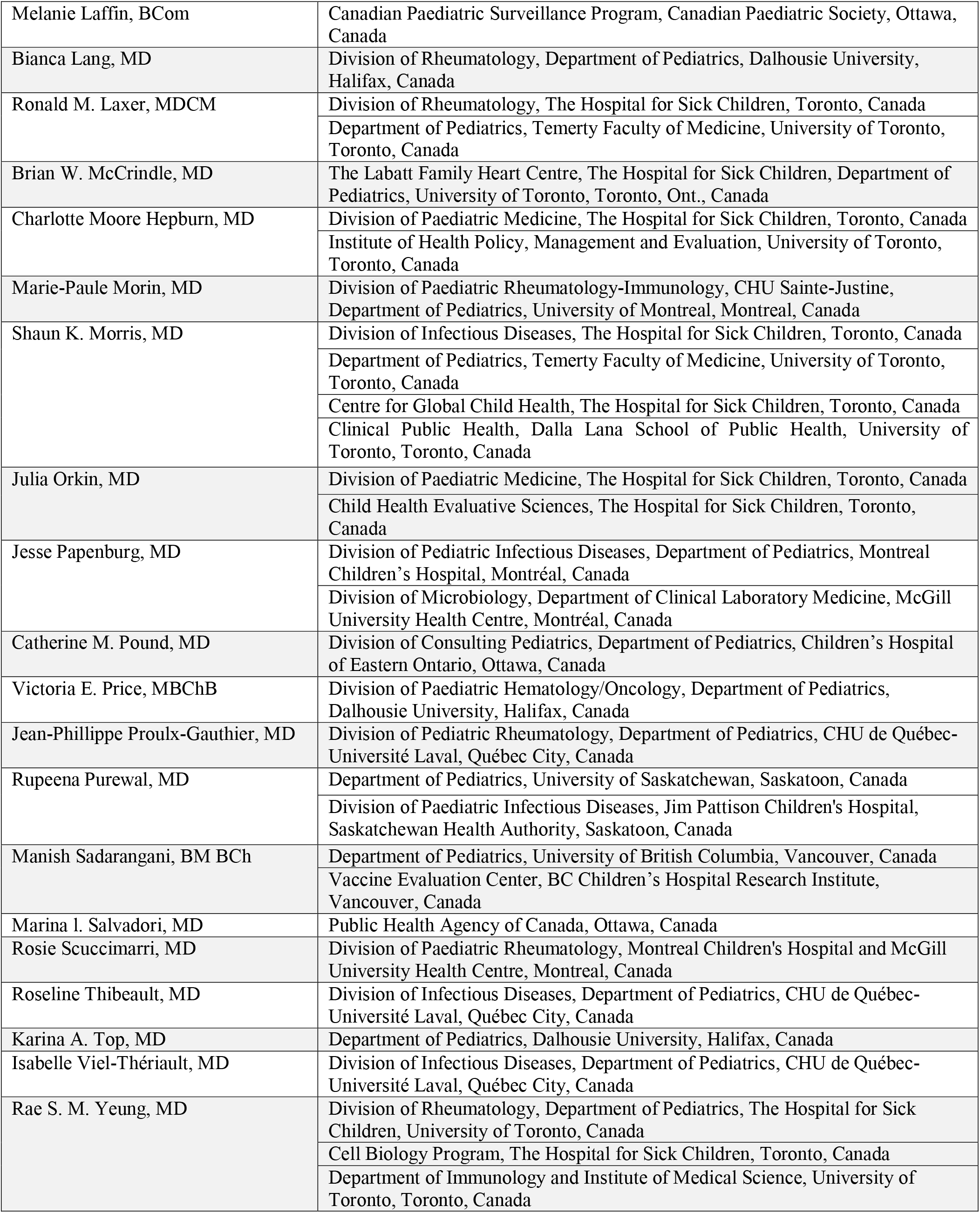

